# Revisiting the link between childhood adversity and stress-sensitive brain regions in psychosis and bipolar disorder: A systematic review and meta-analysis

**DOI:** 10.64898/2026.07.17.26358306

**Authors:** Teya Petrova, Andreas Øy Tennfjord, Daniela Cavero, Aisling Holohan, Melissa Kizilkaya, Ava Ebrahimian-Roodbari, Inès Lepreux, Marie Reimer, Lucia Sideli, Romayne Gadelrab, Giulia Trotta, Victoria Rodriguez, Ole A. Andreassen, Paul Klauser, Luis Alameda, Monica Aas

**Affiliations:** Service of General Psychiatry, Lausanne University Hospital and University of Lausanne, Lausanne, Switzerland; Vestre Viken hospital Trust, Norway; Department of Psychology, The University of Hong Kong, Hong Kong, China; Social, Genetic and Developmental Psychiatry Centre, Institute of Psychiatry, Psychology and Neuroscience, King’s College London, London, UK; Department of Psychosis Studies, Institute of Psychiatry, Psychology & Neuroscience, King’s College London, London, UK; Centre for Precision Psychiatry, Division of Mental Health and Addiction, Oslo University Hospital and University of Oslo, Norway; Department of Human Science, LUMSA University, Rome, Italy; Centre for Psychiatric Neuroscience, Department of Psychiatry, Lausanne University Hospital and University of Lausanne, Lausanne, Switzerland; Division of Child and Adolescent Psychiatry, Department of Psychiatry, Lausanne University Hospital and University of Lausanne, Lausanne, Switzerland; National Psychosis Unit, South London and Maudsley Foundation Trust, National Health Services, London, UK; Instituto de Investigación Sanitaria de Sevilla, IBiS, Hospital Universitario Virgen del Rocío, Universidad de Sevilla, Sevilla, 41013, Spain

## Abstract

**Background:** Brain abnormalities related to childhood adversity (CA) have been reported across clinical presentations in psychotic disorder (PD) and bipolar disorder (BD). This systematic review and meta-analysis examined gray matter volume (GMV) alterations linked to CA in PD and BD.

**Methods:** A PRISMA-compliant systematic review was conducted (PROSPERO ID: CRD42022351133). The EMBASE, MEDLINE, and PsycINFO databases were searched from inception to June 2024 for studies investigating CA and structural brain imaging in PD and BD. Study quality was assessed with the Newcastle Ottawa Scale (NOS). Data were extracted and synthesized accounting for sex differences and CA subtypes with brain findings categorized by the presence and direction of associations. Meta-analyses were performed for hippocampal and amygdala volumes.

**Results:** In the systematic review (*k* = 29), 3,056 participants with PD and BD (mean age = 36.6±16.1; 47% female), published between 2011 and 2023, were included. Study quality was fair, with high heterogeneity. Most studies reported significant negative associations between CA and GMV, especially in prefrontal regions, while findings for the hippocampus and amygdala were largely null or inconsistent. Meta-analyses of a study subset identified no significant association between CA and hemisphere-specific and combined volumes of the hippocampus (*k* = 5; *p* ≥ 0.66) or amygdala (*k* = 4; *p* ≥ 0.87).

**Conclusion:** CA was not consistently associated with hippocampal or amygdala volume alterations in PD and BD. More consistent evidence emerged for reduced GMV in prefrontal regions, suggesting that neurobiological impact of CA may be more robustly captured at the cortical level.

## Introduction

Individuals with a psychotic disorder (PD) and bipolar disorder (BD) report higher rates of childhood adversities (CA) than the general population ^1,2^. Exposure to CA has consistently been linked to greater illness severity ^3,4^ and a broad range of cognitive impairments, including reduced cognitive flexibility, lower IQ, poorer attention, emotion recognition, and working memory deficits ^5,6^. In parallel, neuroimaging evidence suggests that CA is associated with distinct brain volumetric alterations in subcortical, temporal and frontal regions ^7,8^ that may underly these cognitive features. Previous meta-analyses have explored the link between CA and brain alterations in healthy adults ^9,10^. However, the role of CA in brain abnormalities in individuals with PD and BD remains insufficiently understood.

This gap is particularly relevant as increasing evidence conceptualizes PD and BD as part of the same clinical and neurobiological continuum characterized by overlapping genetic, neural, and psychological mechanisms ^11^. Accordingly, examining CA-related brain alterations beyond traditional diagnostic boundaries may be more informative than focusing on disorder-specific effects. In both PD and BD, converging evidence indicates reduction in total gray matter volume (GMV) ^12^, as well as volumetric reductions in the hippocampus ^13^ and the amygdala ^14^, key regions for stress and emotional regulation. Within this transdiagnostic framework, CA may function as a common environmental risk factor that disrupts shared neurodevelopmental trajectories rather than affecting specific diagnostic categories differentially.

It has been hypothesized that individuals exposed to trauma and those within PD and BD may share a common mechanism involving dysregulation of the hypothalamic-pituitary-adrenal (HPA) axis ^15^. Exposure to early life stress is thought to induce prolonged activation of this system, leading to elevated glucocorticoid levels that disproportionally affect stress-sensitive brain regions, such as the hippocampus and amygdala. These regions are highly enriched with glucocorticoid receptors, making them particularly vulnerable to neurotoxic effects of chronic stress exposure. Prolonged glucocorticoid activity may result in dendritic atrophy and reduced synaptic plasticity^16^ providing a mechanistic explanation for observed volumetric reductions and suggesting a potential pathway through which CA may induce a transdiagnostic vulnerability. By disrupting neurodevelopmental trajectories common to both PD and BD, CA leads to convergent structural brain abnormalities (i.e., volume reductions), regardless of traditional clinical labels ^17,18^.

Despite this biological framework, findings regarding CA-related brain alterations remain inconsistent. In the general population, meta-analytic evidence has yielded mixed results. Similar variability has been observed in clinical samples. Earlier structural magnetic resonance imaging (MRI) in psychiatric populations often did not account for trauma history. While earlier structural MRI studies have overlooked the role of trauma, the inclusion of CA measures in clinical samples has expanded substantially in recent studies. This growing body of interest of the literature supports the hypothesis that trauma-related alterations may be involved in the disease etiopathogenesis ^19^.

Previous reviews have examined this question within diagnostic categories. Cancel et al. ^20^ conducted a systematic review examining the link between CA and neuroimaging data in individuals with schizophrenia and found 15 studies reporting evidence of a negative association between CA and GMV decrease, alterations in white matter integrity, and alterations of functional connectivity. Similarly, Zovetti et al. ^21^ reviewed 14 studies focusing on the link between CA and brain alterations in BD and concluded a volume reduction in several brain regions, including the hippocampus, amygdala, thalamus, and frontal cortex.

Building on these findings, the current work provides an updated systematic review of the literature, extending beyond the most recent search conducted in 2018 ^21^, with a substantial amount of research published since. In addition, we conduct a first meta-analysis to quantitatively assess the association between CA and brain volume alterations transdiagnostically across PD and BD.

By focusing on published structural MRI studies, we aim to explore the association between CA and GMV, with a focus on the hippocampus and the amygdala, as these regions have been most frequently examined in this context, but we also include studies that identified other GMV alterations. We conduct a meta-analysis of the association between CA and the volume of these GMV structures, examining various important moderators such as sex and neuroimaging methods, variables that have previously been linked to inconsistent findings ^10^. CA subtypes (e.g., abuse and neglect) are discussed descriptively in the systematic review.

To our knowledge, this is the first systematic review and meta-analysis to include individuals with PD and BD as part of the same neurobiological continuum in addressing this gap in the literature. Within this transdiagnostic framework, we aim to clarify whether CA is associated with (1) reduced hippocampal and/or amygdala volume and (2) broader reductions in GMV across other regions.

## Methods

This systematic review and meta-analysis were pre-registered with PROSPERO International Prospective Register of Systematic Reviews. The protocol ID number is CRD42022351133. PRISMA (Preferred Reporting Items for Systematic Reviews and Meta-Analyses) 2020 guidelines were followed to ensure clear reporting of review findings ^22^; see **Supplementary Table 1**.

### Search Strategy

The primary search was conducted through the following databases: MEDLINE, EMBASE and PsycINFO via the Ovid platform. Medical Subject Headings (MeSH) and keywords related to (i) PD and BD; (ii) childhood adversity; and (iii) brain image data (e.g., structural MRI, GMV) were searched using the Boolean operator “OR” and the three categories were combined using “AND”. A manual search across references of papers that had already been found to meet criteria was also conducted, as well as from final included papers and from relevant similar reviews conducted on the same topic as this review ^20,21^, to identify further studies that may be eligible for inclusion (see **Supplementary Table 2** for the full list of search terms used).

### Eligibility Criteria

According to the PICO framework ^23^, included studies were (1) (P) conducted on adults (aged between 18-80 years) of any gender with a formal diagnosis of a PD or a BD, based on validated diagnostic manuals including current or previous versions of either the Diagnostic and Statistical Manual of Mental Disorders (DSM) ^24^ or the International Classification of Diseases (ICD) ^25^, see **Supplementary Table 3**; (2) (I) reported the presence of CA (see **Operationalization of the predictors section** below); (3) (C) compared brain GMV of people with and without CA amongst individuals with PD or BD, or examined associations using a non-trauma group as reference (4) (O) quantitatively evaluated changes of brain GMV in relation to CA (see section **Operationalization of the outcomes** below); (5) included clinical observational studies, cohort studies, or case-control studies; (6) were originally published in English; (7) were published in peer-reviewed journals.

To be included in the meta-analysis, only studies that (1) reported volume measurements of the hippocampus and/or amygdala; (2) provided corresponding statistical associations (e.g., correlation coefficients or other effect sizes – see details below) with measures of CA were eligible for inclusion.

### Operationalization of the predictors (childhood adversities)

CA was either defined as general adversity (GA, meaning a composite measure broadly defined) or subtypes - sexual abuse (SA), physical abuse (PA), emotional abuse (EA), physical neglect (PN), and emotional neglect (EN). Exposure was considered present if any of these subtypes was assessed by the study. CA subtypes were analyzed only when independent statistical analyses were conducted and reported in the studies.

### Operationalisation of the outcome measures (structural brain volumetric changes)

The primary outcome of interest is volumetric brain differences of GMV in individuals with PD (including schizophrenia, schizoaffective disorder and first-episode psychosis) and BD. Of particular interest are the hippocampus and amygdala volumes, given their established roles in stress response and prior links to CA ^19^.

An expanded category labeled “other GMV”, including also GMV measures from additional brain regions (e.g., prefrontal cortex, thalamus, nucleus accumbens), was also considered for the systematic review if they were reported in either whole-brain or ROI studies.

Studies employing structural MRI techniques (e.g., voxel-based morphometry, manual or automated tracing) were eligible for inclusion. Studies that used multiple neuroimaging modalities remained eligible, but only structural MRI data were extracted and analyzed.

### Screening procedure and data extraction

All studies found from the initial search were exported into an Excel file and were screened based on title and abstract in duplicate by two authors (DC and AT), with any inconsistencies cross-checked by a third author (MA). Following this, full-text screenings of articles were conducted using the same procedure; any discrepancies were resolved through consensus. The following information was extracted in duplicate from each relevant study: surname of first author, year and country of publication, sample size, mean age, percentage of female participants, study design, clinical diagnoses, CA/subtype, and instrument used to measure CA, technique used to measure volumetric data (manual or automated segmentation), brain region of interest (ROI), brain regions reported to be affected by CA, whether analyses were adjusted and by which confounders, and lastly, the association (effect size, direction with respective test and significance) between the reported CA (predictor) and the reported volumetric structural brain imaging data (outcome).

### Qualitative data analysis

For the systematic review, we appraised the evidence by classifying each association as either significant or not significant, based on a statistical threshold (p < 0.05 and 95% confidence interval included the null value). Then, among the significant findings, associations were further categorized as either negative (more CA was associated with smaller brain volumes) or positive (more CA was associated with greater brain volumes). The proportions of significant positive and negative associations were then calculated across studies as independent analyses; mixed results were included in both categories. Results from studies examining trauma subtypes or sex differences were included in the overall count of CA findings when they reported associations with brain volume. Some studies that used a whole-brain approach also reported ROI results of the hippocampus and amygdala ^26–28^ and were therefore included in these counts. When studies used whole-brain analyses without reporting region-specific (hippocampal or amygdala) results, these regions were not classified as non-significant and were excluded from the corresponding ROI-based counts.

### Quantitative data analysis

Statistical analyses were conducted using Comprehensive Meta-Analysis (CMA-version-4) ^29^. Effect sizes reflecting the association between CA and brain volumes were reported as Pearson’s r. In studies where alternative statistics were reported, such as standardized β, Cohen’s d, or F-values, these were converted to Pearson’s r using established methods to ensure comparability as previously described ^30^ and used in previous meta-analyses ^3^. The software computed correlation coefficients based on the extracted coefficients. These were first transformed using Fisher’s r-to-z transformation to reduce bias, particularly important when synthesizing data from a limited number of studies, and then converted back to r values for interpretation ^31^.

The primary analysis focused on the pooled effect sizes of the association between CA and left and right hippocampal and amygdala volumes given the well-established hemispheric asymmetry ^32^ to ensure that lateralized effects were not overlooked. Secondary analyses were performed on combined total hippocampal and amygdala volume, as some studies reported them as such ^33,34^. For these secondary analyses, studies that reported separate associations between CA and left and right hemispheric volumes, but not with total, a combined association was computed to obtain a single effect size per region in relation to CA. This was done using inverse-variance-weighted averaging of Fisher’s z-transformed correlation coefficients, which were then back-transformed to Pearson’s r ^35^. Studies reporting subfield volumes ^36–38^ or presenting sex-stratified results ^39^ were not included in the meta-analysis. In the primary analyses, only overall CA coefficients from patient samples were included. Studies reporting associations in combined patient and control samples were excluded from the meta-analysis. For the secondary analyses of total hippocampal and amygdala volume, one study reported a coefficient specific to emotional neglect; this value was included to maximize data coverage, while all other studies contributed general CA measures.

The presence of between-study heterogeneity was assessed with the Q-test (p < 0.05 indicating potential heterogeneity) and the magnitude of heterogeneity with the I^2^ (I^2^ > 50 % is considered high heterogeneity) ^40^. High heterogeneity was expected; therefore, random-effects meta-analyses were conducted. Only one meta-analysis per study sample was conducted to avoid double counting bias. Meta-analyses were conducted when there were at least four samples per meta-analysis in the same volumetric brain region. All p-values reported in the meta-analysis were two-sided, with a significance level of p < 0.05, with 95% confidence intervals.

To explore potential sources of heterogeneity and the moderating effect of important factors, meta-regression analyses were conducted using age, sex, illness stage (first-episode vs. chronic illness), segmentation method (manual vs. automated), and study quality (e.g., NOS). Finally, to address diagnostic heterogeneity, a post-hoc sensitivity analysis was conducted by restricting dataset to only chronic illness samples and excluding first-episode cohorts.

### Quality assessment and risk of bias

The quality assessment of the included studies was conducted using a revised version of the Newcastle-Ottawa Scale (NOS) ^41^ for non-randomized cohort studies by two reviewers (DC and AT). Any discrepancies were resolved through consensus and input from a third reviewer (MA). Papers were assessed using the NOS ‘star system’ and judged on three categories: selection, comparability, and outcome. Finally, each paper was given a total quality rate. See **Supplementary Tables 4, 4a** for more details.

To assess publication bias, referring to the systematic overrepresentation of studies with statistically significant or expected results in the published literature, we looked for asymmetry in the funnel plots and conducted Egger’s test (p < 0.05 indicates potential publication bias) ^42^.

## Results

### Summary of Search

The initial screening of the 1635 studies originally identified in the first search (May 2022) (DC and AT) identified 24 studies meeting inclusion criteria, involving 2,062 participants. A subsequent updated search of the literature (June 2024) (TP and LA) yielded an additional 5 studies, adding 994 more participants, resulting in a total sample of 3,056, with a mean age of 36.6±16.1. The sample was 47% female. Details of the screening process are reported in the PRISMA flow chart in **Fig. 1**.

**Fig. 1.**
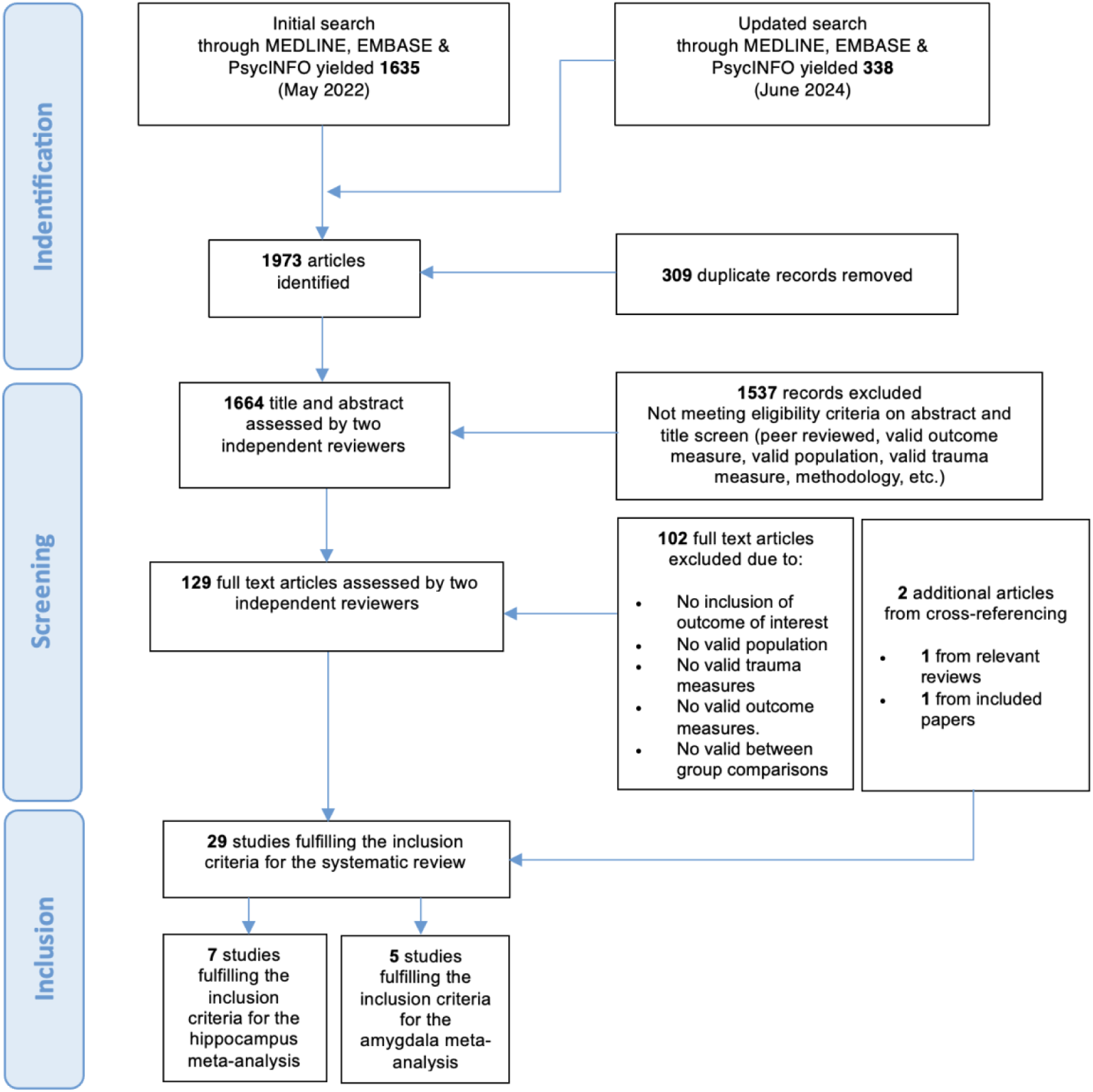
Prisma Flowchart of database search by June 2024

The sample sizes of eligible studies ranged from 20 to 788. Overall, the DSM, Fourth Edition (DSM-IV) was the most used diagnostic instrument (83%). Almost half of the studies (48%) were published in the last 5 years (from 2019 to 2024). Most studies were conducted in Europe or North America, with six studies conducted in non-Western countries (China, South Africa, Brazil and Korea). As for the CA outcome measure, 65% of the studies used the Childhood Trauma Questionnaire (CTQ) ^43^, most commonly using the composite score for the analyses, and a smaller number applied a binary classification of trauma exposure (see **Table 1** and **Supplementary Table 5** for full details of CA used measures). The quality of included studies ranging from 4 (poor) to 6 (good) based on the Newcastle-Ottawa Scale (NOS). More specifically, 45% of the included studies had a quality rating of “good” (score of 6) and 52% of “fair” rating (score of 5). Full details are provided in **Supplementary Table 4 and 4a**.

**Table 1.** Summary of the results: associations between CA and GMV alterations.

| Study,<br>(Country) | Sample size<br>(% female<br>patients) | Clinical<br>Diagnosis | Mean age<br>(±SD) | CA<br>Measure<br>(Subtypes) | Neuroimaging<br>method | Reported brain region volumes in<br>relation to CA | Reported significant associations<br>with CA and directionality |
| --- | --- | --- | --- | --- | --- | --- | --- |
| Aas et al. <sup>45</sup><br>(UK) | 45<br>(37%) | FEP | 27.4<br>(± 7.9) | CTQ total<br>score | ROI, manual<br>segmentation | Amygdala, hippocampus | ↓ Total amygdala volume |
| Alameda et al. <sup>53</sup><br>(Switzerland) | 64<br>(36%) | FEP | 25.4<br>(± 5.5) | CTQ total<br>score | ROI, automated<br>segmentation | Amygdala, hippocampus | ↓ Left hippocampal volume |
| Armio et al. <sup>58</sup><br>(Finland) | 75<br>(45%) | FEP | 26.8<br>(± 6.0) | TADS total<br>score | ROI, automated<br>segmentation | Amygdala subfields | ↓ Amygdala lateral nucleus<br>volume |
| Begemann et al.<br><sup>26</sup><br>(Holland) | 327<br>(44%) | SCZ<br>BP-I | 44.4<br>(± 13.7) | CTQ total<br>score and<br>subtypes<br>(SA, PA, EA,<br>PN, EN) | Whole-brain and<br>ROI, automated<br>segmentation | 68 grouped cortical regions and<br>also ROIs: hippocampus, amygdala,<br>nucleus accumbens, caudate,<br>pallidum, putamen, thalamus | None in PD<br>↓ Frontal lobe in BD |
| Bendetti et al. <sup>44</sup><br>(Italy) | 20<br>(30%) | SCZ | 33.2<br>(± 7.6) | RFQ total<br>score | ROI, VBM | ACC, PFC, amygdala, hippocampus | ↑ ACC volume<br>↑ PFC volume |
| Cancel et al. <sup>59</sup><br>(France) | 21<br>(29%) | SCZ | 32.1<br>(± 8.3) | CTQ total<br>score and<br>subtypes<br>(SA, PA, EA,<br>PN, EN) | Whole-brain,<br>VBM | | ↓ total GMV associated with EN<br>↓ DLPFC |
| Cao et al. <sup>62</sup><br>(China) | 150<br>(40%) | BD-II | 22.8<br>(± 7.2) | CTQ total<br>score and<br>subtypes<br>(SA, PA, EA,<br>PN, EN) | Whole-brain,<br>VBM, SBM | | ↓ Right middle frontal gyrus<br>volume associated with SA<br>↑ Right orbital middle frontal<br>gyrus volume associated with PN |
| Colic et al. <sup>39</sup><br>(USA) | 119<br>(59%) | BD | 31.1<br>(± 12.0) | CTQ total<br>score | ROI, automated<br>segmentation | Hippocampus, PFC regions | ↓ Left hippocampus (in women)<br>↓ Right superior frontal surface<br>area (in men) |
| Del Re et al. <sup>37</sup><br>(USA) | 788<br>(58%) | SCZ,<br>SZA<br>BD-I | 38.2<br>(± 22.4) | CTQ total<br>score | ROI, automated<br>segmentation | Hippocampus | ↓ Right anterior hippocampus<br>↓ Left posterior hippocampus<br>↓ Right posterior hippocampus |
| Duarte et al. <sup>27</sup><br>(Brazil) | 39<br>(59%) | BD-I | 41.7<br>(± 12.0) | CTQ total<br>score and<br>subtypes<br>(SA, PA, EA,<br>PN, EN) | Whole-brain,<br>VBM | ROIs: hippocampus, amygdala,<br>thalamus | ↓ GMV in right DLPFC associated<br>with PA<br>↓ Bilateral thalamus associated<br>with PN and EN |
| Du Plessis et al. <sup>38</sup><br>(South Africa) | 79<br>(26%) | FEP | 23.0<br>(± 7.0) | CTQ total<br>score | ROI, automated<br>segmentation | Hippocampal subfields | None |
| Elvsåshagen et. al.<br><sup>36</sup><br>(Norway) | 37<br>(70%) | BD-II | 33.4<br>(± 6.6) | CTQ total<br>score | ROI | Hippocampus and subfields | None |
| Fan et al. <sup>34</sup><br>(China) | 127<br>(43%) | SCZ | 40.8<br>(± 13.1) | CTQ total<br>score and<br>subtypes<br>(SA, PA, EA,<br>PN, EN) | ROI, automated<br>segmentation | Total GMV, thalamus, caudate,<br>putamen, pallidum, nucleus<br>accumbens, amygdala,<br>hippocampus | ↓ Amygdala volume associated<br>with EN |
| Frissen et al. <sup>61</sup><br>(The Netherlands) | 89<br>(33%) | SCZ | 28.1<br>(± 7.0) | CTQ total<br>score | ROI, automated<br>segmentation | Total GMV | ↓ total GMV |
| Harmata et al. <sup>63</sup><br>(USA) | 131<br>(40%) | BD-I | 39.8<br>(± 13.4) | ACE score | ROI | Cerebellum | ↓ Cerebellar volume |
| Hernaus et al. <sup>47</sup><br>(Belgium) | 89<br>(40%) | SCZ | 28.1<br>(± 7.0) | CTQ total<br>score | ROI, automated<br>segmentation | Hippocampal volume | None |
| Hoffmann et al. <sup>48</sup><br>(Australia) | 153<br>(29%) | SCZ<br>SZA | 38.2<br>(± 10.1) | CAQ,<br>(SA, PA, EA,<br>EN) | ROI, automated<br>segmentation | Caudate, putamen, nucleus<br>accumbens, DLPFC, hippocampus,<br>ICV | ↓ Caudate volume associated<br>with PA |
| Hoy et al. <sup>54</sup><br>(UK) | 21<br>(57%) | FEP | 31.9<br>(± 10.4) | TEC total<br>score | ROI, manual<br>segmentation | Amygdala, hippocampus | ↓ Total amygdala volume<br>↓ Left hippocampal volume |
| Janiri et al. <sup>56</sup><br>(Italy) | 105<br>(50%) | BD-I<br>BD-II | 44.1<br>($\pm 12.1$) | CTQ total<br>score | ROI, automated<br>segmentation | Amygdala, hippocampus, nucleus<br>accumbens, caudate, pallidum,<br>putamen, thalamus | ↑ Amygdala volume<br>↑ Hippocampal volumes |
| Janiri et al. <sup>57</sup><br>(Italy) | 104<br>(50%) | BD-I<br>BD-II | 44.0<br>($\pm 12.2$) | CTQ total<br>score | ROI, automated<br>segmentation | Hippocampus and subfields | Hippocampus subfields:<br>↑ subiculum<br>↑ presubiculum<br>↑ CA1 |
| Millman et al. <sup>49</sup><br>(USA) | 41<br>(51%) | SCZ | 33.7<br>($\pm 7.2$) | CTQ total<br>score | ROI, automated<br>segmentation | Amygdala, hippocampus | ↑ Left amygdala volume |
| Poletti et al. <sup>55</sup><br>(Italy) | 86<br>(70%) | BD-I | 45.4<br>($\pm 12.8$) | RFQ total<br>score | ROI | Hippocampus | ↓ Hippocampal volume |
| Quidé et al. <sup>60</sup><br>(Australia) | 107<br>(45%) | SCZ<br>SZA<br>BD-I | 39.4<br>($\pm 12.2$) | CTQ total<br>score | Whole-brain,<br>SBM | | In PD :<br>↓ PCC/precuneus<br>↓ Parietal lobule<br>↓ Postcentral gyrus<br>↑ Left middle temporal gyrus<br>In BD : none |
| Rokita et al. <sup>50</sup><br>(Ireland) | 46<br>(26%) | SCZ<br>SZA | 43.8<br>($\pm 10.9$) | CTQ total<br>score and<br>subtypes<br>(SA, PA, EA,<br>PN, EN) | ROI, automated<br>segmentation | Amygdala, hippocampus, ACC | None |
| Ruby et al. <sup>33</sup><br>(USA) | 28<br>(29%) | SCZ | 31.5<br>($\pm 9.7$) | Early Trauma<br>Inventory | Whole-brain,<br>automated<br>segmentation;<br>ROI, manual<br>segmentation | Amygdala, hippocampus, total<br>GMV | ↑ Total amygdala volume |
| Sheffield et al. <sup>51</sup><br>(USA) | 60<br>(47%) | SCZ<br>SZA<br>BP-I | 35.5<br>($\pm 13.3$) | CTQ total<br>score and<br>subtypes<br>(SA, PA, EA,<br>PN, EN) | ROI | Total GMV, hippocampus,<br>amygdala, PFC (7 regions) | ↓ total GMV associated with SA<br>↓ Left Middle Frontal Gyrus |
| Song et al. <sup>28</sup><br>(South Korea) | 36<br>(56%) | BD-I<br>BD-II | 30.6<br>($\pm 5.3$) | CTQ total<br>score | Whole-brain | | ↓ Right precentral gyrus volume |
| Souza-Queiroz et al. <sup>52</sup><br>(France) | 32<br>(38%) | BD | 35.8<br>(± 11.2) | CTQ total score and subtypes<br>(SA, PA, EA, PN, EN) | ROI | Amygdala, hippocampus | None |
| Tseng et al. <sup>64</sup><br>(Taiwan) | 37<br>(57%) | SCZ | 35.7<br>(± 9.3) | BBTS total score | Whole-brain |  | None |

Abbreviations: ACC (Anterior Cingulate Cortex), ACE (Adverse Childhood Experiences), BBTS (Brief Betrayal Trauma Survey), BD-I (Bipolar disorder type I), BD-II (Bipolar disorder type II), CAQ (Childhood Adversity Questionnaire), CTQ (Childhood Trauma Questionnaire), DLPFC (Dorsolateral Prefrontal Cortex), EA (Emotional Abuse), EN (Emotional Neglect), GMV (Gray Matter Volume), PA (Physical Abuse), PFC (Prefrontal Cortex), PN (Physical Neglect), RFQ (Risky Families Questionnaire), ROI (Region of Interest), SA (Sexual Abuse), SBM (Surface-Based Morphometry), SCZ (Schizophrenia), SZA (Schizoaffective disorder), TADS (Trauma and Distress Scale), VBM (Voxel-Based Morphometry)
Note: CA subtype results are specified only where independent statistical analyses were performed. “↓” indicates a negative association. “↑” indicates a positive association

The included studies used different neuroimaging methods to test for the association between CA and brain volumes. Overall, 76% used a hypothesis-driven, region of interest (ROI) approach to investigate abnormalities previously identified in MRI studies of the impact of CA on PD and BD, most commonly the hippocampus and the amygdala. Among the ROI studies, the vast majority (81%) used an automated segmentation approach (e.g., FreeSurfer), rather than manual tracing (14%). One study used voxel-based morphometry (VBM) ^44^. The remaining 24% of the studies had a whole-brain approach, primarily using VBM (70%), followed by source-based morphometry (SBM) (20%), and automated segmentation of the entire brain (10%).

Regarding diagnostic distribution across the systematic review, 28% of the studies included individuals diagnosed with schizophrenia (k=8), 17% with first-episode psychosis (k=5), 34% with bipolar disorder (k=10), and 21% were composed of individuals with different diagnostic labels (k=6).

### Association between childhood adversity and brain volumes in PD and BD

The narrative findings are presented below based on regional anatomical categories. The results from each study are summarized in **Table 1**.

Across the 29 studies investigating the impact of CA on brain volume in individuals with PD and BD, 79% (23/29) of studies reported at least one significant association, while 21% (6/29) reported only null findings. Among the studies indicating significant results, 38 independent analyses were reported, of which 71 % (27/38) indicated a negative relationship between CA and brain volume. In contrast, 29% (11/38) indicated a positive associations. Detailed findings for specific brain regions are provided below (see **Table 1**; for more information, see **Supplementary Table 5**).

### Hippocampus

Twenty-one studies investigated the association between CA and hippocampal volume. Among them, the majority 67% (14/21) did not report significant findings ^27,33,34,36,38,44–52^. In contrast, 33% (7/21) reported significant associations. Among these seven studies, 11 independent analyses were performed, of which 64% (7/11) supported a negative association with CA ^37,39,53–55^ and 36% (4/11) supported a positive association ^56,57^.

### Amygdala

Fourteen studies examined amygdala volume in relation to CA, among which 50% (7/14) reported no association ^27,44,46,50–53^ and 50% (7/14) identified significant effects. Of the seven independent analyses, 57% (4/7) identified a negative association ^34,45,54,58^ and 43% (3/7) a positive association ^33,49,56^ between CA and amygdala volume.

### Other GMV regions

Across seventeen studies assessing the impact of CA on GMV in additional brain regions (e.g., frontal lobe regions, ACC, caudate and nucleus accumbens) using both ROI and whole-brain approaches 71% (12/17) showed significant associations and only 29% (5/17) reported null findings. Across the twelve studies reporting significant findings, a total of 20 independent analyses were conducted. Here, a majority of 80% (16/20) showed negative associations ^27,28,39,46,48,51,59–63^, and only 20% (4/20) showed positive associations ^44,60,62^. This suggests more consistent evidence for CA-related reductions in GMV outside the hippocampus and amygdala.

In summary, these findings reveal a lack of consensus regarding the association between hippocampal volume and CA. Results for the amygdala are similarly inconsistent, showing divergent directionality of the association, with some findings demonstrating volume reductions and others toward increases, preluding any definitive conclusion regarding directionality. In contrast, within other GMV regions (e.g., PFC, precuneus, caudate, thalamus), a greater proportion of studies observed negative associations with CA. Five studies^34,44,46,59,62^ using whole-brain approach consistently identified significant associations within prefrontal regions (DLPFC, ACC, vmPFC, frontal gyrus), suggesting that these areas may more consistently reflect the impact of early-life adversity in PD and BD.

### Subgroup analyses

#### Subtypes of Trauma

A total of nine studies explored the association between specific CA subtypes and regional brain volumes. Findings regarding physical abuse were characterized by negative associations within the caudate ^65^ and DLPFC ^27^. Similarly, Sheffield et al. ^51^ and Cao et al. ^62^ found negative correlations between SA and total GMV and frontal gyrus volume, respectively. Investigations into neglect revealed divergent results. Cancel et al. ^59^ reported a negative association between EN and total GMV and Duarte et al. ^27^ reported a negative correlation between thalamic volume and EN and PN. Similarly, Fan et al. ^66^ found a reduction in total amygdala volume linked to EN. In contrast, the study by Cao et al. ^62^ identified a positive association between PN and the frontal gyrus volume. Finally, three studies that explored the impact of CA subtypes did not yield significant results ^46,50,52^, highlighting a persistent heterogeneity in how distinct trauma types manifest neurobiologically in PD and BD.

#### Sex differences

Only a few studies examined sex-specific effects. Colic et al. ^39^ reported a negative association between CA and hippocampal volume in female participants, whereas in male participants, CA was negatively associated with frontal pole surface area. Du Plessis et al. ^38^ found a positive association between CA and hippocampal fissure volume in female patients, an effect not observed in either male patients or healthy controls. Ruby et al. ^33^ found associations exclusively in men, specifically a negative correlation between CA and both whole-brain volume and amygdala-to-whole brain ratio. Conversely, Milman et al. ^49^ did not support sex differences. These findings underscore the need for more investigations into sex-specific neurobiological responses to CA.

### Meta-analysis

Quantitative meta-analyses were restricted to the hippocampus and amygdala due to data availability. To ensure statistical validity and account for within-subject correlation, we prioritized hemisphere-specific analyses (left and right) for the hippocampus and amygdala. As two studies ^33,34^ reported effect sizes of combined right and left volumes, we also conducted two exploratory meta-analyses to ensure no data was excluded from the synthesis (see **Table 2**). Forest plots can be found in the **Supplementary Material** (**Figures 1-3**).

**Table 2.**
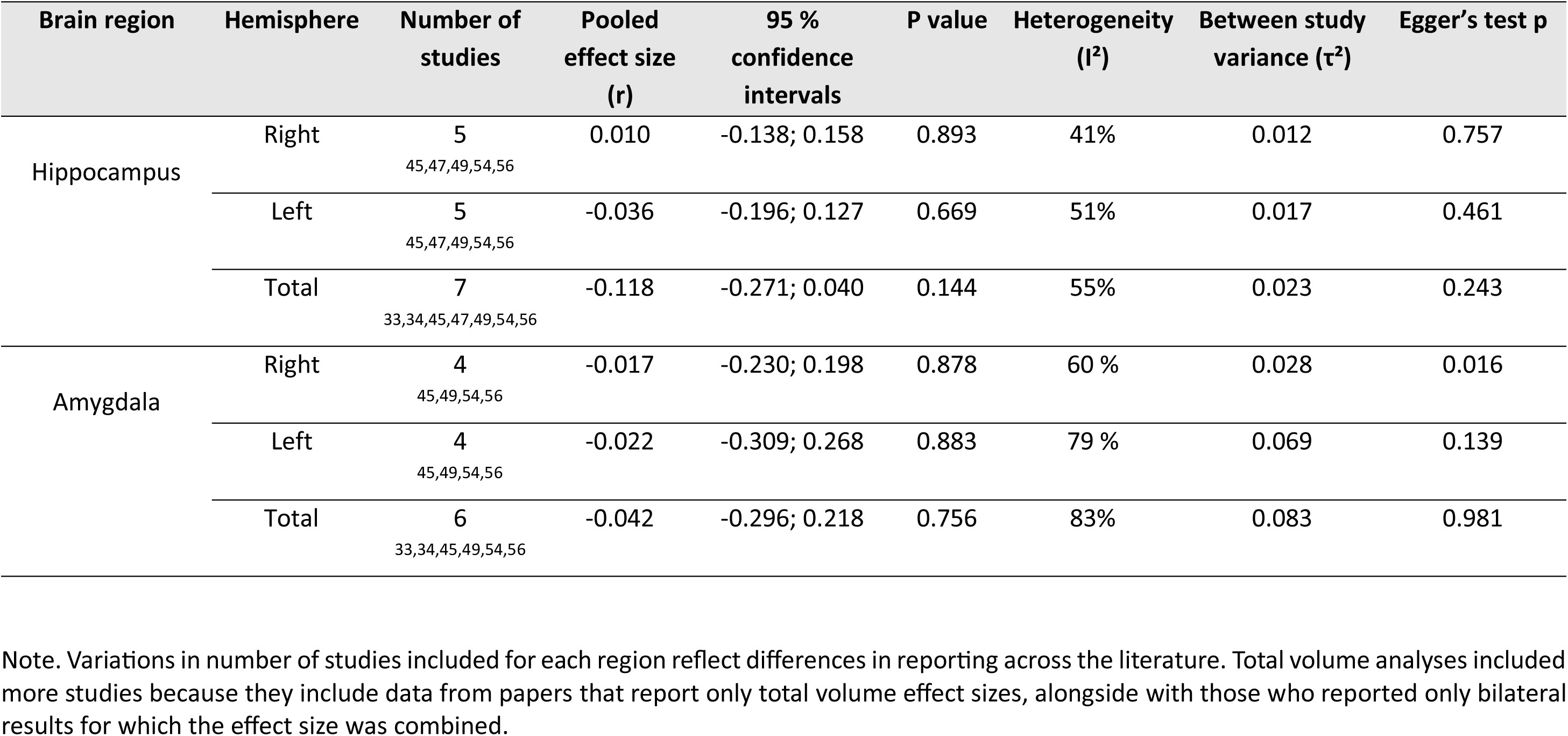
Meta-analysis results.

| Brain region | Hemisphere | Number of studies | Pooled effect size (r) | 95 % confidence intervals | P value | Heterogeneity (I<sup>2</sup>) | Between study variance ($\tau^2$) | Egger's test p |
| --- | --- | --- | --- | --- | --- | --- | --- | --- |
| Hippocampus | Right | 5<br>45,47,49,54,56 | 0.010 | -0.138; 0.158 | 0.893 | 41% | 0.012 | 0.757 |
|  | Left | 5<br>45,47,49,54,56 | -0.036 | -0.196; 0.127 | 0.669 | 51% | 0.017 | 0.461 |
|  | Total | 7<br>33,34,45,47,49,54,56 | -0.118 | -0.271; 0.040 | 0.144 | 55% | 0.023 | 0.243 |
| Amygdala | Right | 4<br>45,49,54,56 | -0.017 | -0.230; 0.198 | 0.878 | 60 % | 0.028 | 0.016 |
|  | Left | 4<br>45,49,54,56 | -0.022 | -0.309; 0.268 | 0.883 | 79 % | 0.069 | 0.139 |
|  | Total | 6<br>33,34,45,49,54,56 | -0.042 | -0.296; 0.218 | 0.756 | 83% | 0.083 | 0.981 |

Note. Variations in number of studies included for each region reflect differences in reporting across the literature. Total volume analyses included more studies because they include data from papers that report only total volume effect sizes, alongside with those who reported only bilateral results for which the effect size was combined.

#### Hippocampus

Hemisphere-specific results showed no significant association with CA for either the right (*r* = 0.010, *p* = 0.893) or the left (*r* = −0.036, *p* = 0.669), based on five studies. A secondary analysis of combined hippocampal volume similarly yielded non-significant results (*r* = −0.118, *p* = 0.144).

#### Amygdala

Lateralized analyses for the amygdala reported no significant associations for the right (*r* = −0.017, *p* = 0.878) or the left (*r* = −0.022, *p* = 0.883) hemispheres. Analyses of combined volume also remained non-significant (*r* = −0.042, *p* = 0.756).

#### Heterogeneity and publication bias assessments

Heterogeneity varied across the primary hemisphere-specific analyses, ranging from low in the right hippocampus (*I²* = 41%) to substantial in the left amygdala (*I²* = 79%), as detailed in **Table 2**. While Egger’s regression tests indicated no significant funnel plot asymmetry for either hippocampal hemisphere or the left amygdala, a significant result was observed for the right amygdala (*p* = 0.016). This suggests that the null finding for the right amygdala may be influenced by small-study effects or publication bias and should be interpreted with caution.

#### Meta-regression results

Heterogeneity was further explored through meta-regression models considering age, sex, illness stage (first-episode vs. chronic illness), segmentation method and study quality (NOS scores) as potential moderators. For the hippocampal hemispheres, study quality emerged as a significant moderator only for the right hippocampus (*β* = 0.242, *p* = 0.030), while sex was a significant moderator in the exploratory total volume model (*β* = 0.020, *p* = 0.007) but not for individual hemispheres. Within the amygdala meta-analysis, illness stage, segmentation method, and study quality all appeared as significant moderators for both left and right hemispheres, as well as total combined volume (*p*-values ranging from 0.007 to 0.014). Full details for all meta-regression parameters are provided in **Supplementary Table 6**. Additionally, post-hoc sensitivity analysis isolated strictly to chronic schizophrenia cohorts for amygdala volume (k=4) yielded no significant association between CA and volumetric changes.

## Discussion

This systematic review and meta-analysis provide an updated and quantitatively integrated synthesis of studies examining the relationship between CA and GMV in PD and BD with a focus on the hippocampus and amygdala. Compared to previous reviews ^20,21^, this study advances the field by incorporating a meta-analysis and by jointly examining individuals with PD and BD within the same neurodevelopmental continuum.

The meta-analysis identified no significant volumetric association between CA and hippocampal or amygdala, regardless of hemispheric lateralization. These results challenge the classical neurobiological model of stress, which posits that these limbic structures are the primary targets of early-life adversity. Instead, our systematic review revealed a more consistent pattern of evidence in prefrontal regions (71% of studies showing a negative association), suggesting that the transdiagnostic impact of CA may be more reliably captured in higher-order cortical regions involved in cognitive and emotional regulation rather than in traditional limbic targets. Importantly, these prefrontal findings may be particularly robust, as they frequently emerge from whole-brain analyses rather than hypothesis-driven ROI approaches. Unlike hippocampal findings, which are often tested *a priori* using ROI masks, prefrontal alterations have been identified in studies employing unbiased voxel-based methods, where they survive stringent multiple-comparisons correction ^46,59,62,67^. This suggests that the effect size of CA-related alterations in prefrontal regions may be sufficiently strong and spatially distributed to be detected without anatomical restriction.

These findings call into question the models that prioritize limbic vulnerability as the primary neurobiological consequence of CA, as our results do not provide consistent evidence for volumetric alterations in the hippocampus or amygdala despite their theoretical sensitivity to stress-induced neuroanatomical alterations. Beyond the methodological challenges raised below, a potential explanation is that prefrontal regions may be particularly sensitive to the long-term developmental effects of CA. The prefrontal cortex is among the latest brain regions to fully mature, undergoing prolonged synaptic pruning and myelination into early adulthood ^68,69^. This extended developmental window likely renders it especially vulnerable to chronic stress exposure, potentially leading to altered maturation trajectories. In this context, CA-related reductions in prefrontal GMV may reflect alterations in normative developmental processes such as synaptic pruning and cortical thinning, potentially leading to long-term changes in circuits involved in cognitive and emotional regulation.

### Sources of heterogeneity and moderating factors

In line with previous studies ^10,21,70^, there was high heterogeneity across the included studies which may underlie the absence of significant associations. This heterogeneity appears to arise from multiple sources such as methodological variability (e.g., segmentation techniques, ROI versus whole-brain approaches, covariate adjustment), sample characteristics (e.g., sex distribution, illness stage), clinical variables (e.g., medication use), and differences in trauma assessment. Unlike in non-clinical samples, some of these clinical factors (i.e., medication use, illness stage) introduce substantial neurobiological interference that may obscure the primary signature of CA. Critically, while we adopted a transdiagnostic approach considering psychosis as the common feature, we cannot exclude the possibility that specific diagnoses within the spectrum (e.g., affective vs. non-affective psychotic disorders) exhibit distinc neuroimaging profiles following exposure to CA. To partially account for these potential subgroup differences, we evaluated illness stage (first-episode vs. chronic states) as a moderator in our meta-regression framework and conducted post-hoc sensitivity analysis restricted to exclusively chronic schizophrenia cohorts. In addition, the inclusion of both cohort and case-control designs introduced disparate methodologies into the quantitative analysis. Because cohort and observational designs generally offer a lower baseline level of evidence compared to strictly matches case-control frameworks, their varying susceptibility to selection bias across different clinical settings likely undermined the overall reliability of the pooled meta-analysis results and further emphasized the observed heterogeneity.

Meta-regression analyses were conducted to account for the factors mentioned above, revealing that several variables moderated the association between GMV and CA, with sex emerging as a particularly important factor. Studies including fewer female patients showed stronger negative associations, suggesting that the CA-hippocampal volume relationship may be more pronounced in men and diluted in sex-matched samples or in samples with higher proportions of women. When examining this in detail, only two studies in the meta-analyses of the hippocampus that reported a positive association were matched by sex ^49,56^(including 50% and 51.2% of female participants respectively), while the other five ^33,34,45,47,54^ included predominantly men (median prevalence of 40% females) and one of them did not adjust by sex ^33^. These findings position sex not simply as a confounder but as a key moderator of neurobiological response to CA. In line with this, a previous meta-analysis in a non-clinical sample by Calem et al.^9^ reported significant associations between CA and hippocampal and amygdala volumes, which were no longer observed after adjusting for sex. Indeed, evidence from adults with a history of childhood abuse suggests sex-specific neuroendocrine and corticolimbic differences involving the amygdala, hippocampus and PFC, may contribute to distinct neural trajectories following early adversity^71^. These observations highlight the need for systematically incorporating sex-specific analyses in future research.

Furthermore, heterogeneity was explained by the significant moderating effect of illness stage on amygdala volume, suggesting that associations between CA and volume vary significantly as the disorder progresses from early to chronic stage. While the limited meta-analytic sample did not allow a formal stratification by individual diagnostic labels, illness stage as moderator allowed to evaluate the clinical distinction between the early stages of psychosis compared to chronic presentations (e.g., schizophrenia). This suggests that early stress-related changes may evolve over time, rather than being static. The inconsistencies found could also be explained by the allostatic load model, which posits that high levels of stress initially increase the volume of the amygdala, but extreme or chronic levels of stress may result in a smaller volume ^72^. More specifically, it has been shown that following early life stress the amygdala may expand its volume due to higher dendritic arborization as a compensatory effect after higher functional reactivity; however over time the expansion may transition into toxic damage and atrophy ^72^. Such dynamic trajectories may not be captured in cross-sectional designs. Also, pooling together samples (e.g., first-episode and chronic), which is common in systematic reviews and meta-analysis may further obscure stage-specific effects and dilute associations.

Medication effects further contribute to variability. Lithium, for instance, is known for its neuroprotective effects and is associated with increased GMV ^73^ specifically within the hippocampus and amygdala. While lithium treatment is typically initiated years after the occurrence of CA, it may modify the long-term trajectories of the brain alterations caused by early-life trauma. By promoting neuroplasticity ^74^ or reversing volume loss ^75^ after a disorder has been diagnosed, lithium can act as a compensatory factor to the atrophy associated with CA. Consequently, the inconsistent inclusion of lithium as a covariate likely introduces systematic bias, potentially masking true associations in treated populations. Indeed, in some studies, findings were attenuated after adjusting for medication ^26,27^ and other clinical factors such as duration of illness ^59^.

Another important source of variability concerns the analytical approaches used. Although many studies conducted whole-brain analyses, most relied on hypothesis-driven ROI methods. While ROI approaches are more sensitive due to fewer statistical comparisons, they can also bias findings by restricting the scope of analysis and thus missing global patterns. This methodological divergence is particularly relevant when interpreting limbic versus prefrontal findings, as hippocampal and amygdala volumes are often examined using ROI-based approaches, whereas prefrontal alterations are more consistently identified in whole-brain analyses. This may partially explain why more robust associations were observed in whole-brain studies examining frontal regions compared to limbic-focused ROI studies.

Furthermore, the characteristics of CA instruments used likely influence neurobiological findings. Although most studies used self-reported measures such as the CTQ, these tools are vulnerable to recall bias and usually do not capture important features such as trauma timing. Developmental timing is critical, as early versus later trauma can have distinct neural impacts ^76^, and this was rarely examined in the papers included. Moreover, trauma subtypes (e.g., abuse involving intrusive hostility vs. neglect involving enduring deprivation) have been shown to affect different brain regions ^76^. Specifically, some studies suggest that amygdala volume may be increased in individuals who have experienced neglect but reduced in those who experienced abuse ^77^. Also, abuse more commonly alters threat-processing circuits, whereas neglect is associated with deficits in cognitive and sensory circuits ^78^. The limited and inconsistent assessment of trauma subtypes has prevented the identification trauma-specific signatures in PD and BD given the heterogeneity of measures.

Taken together, the pattern of more consistent findings in the cortical prefrontal regions ^27,28,44,51,60,62^ aligns with previous meta-analyses ^10,20^. Importantly, these alterations appear across schizophrenia, bipolar disorder, and first-episode psychosis samples, supporting the notion that prefrontal GMV reductions may represent a transdiagnostic marker of CA rather than a disorder-specific feature. This is consistent with evidence linking trauma exposure to impairments in executive functioning and emotion regulation across diagnostic categories ^76^. Our qualitative results suggest that the effects of CA on brain structure are unlikely to be localized ^26,36,38,47,50,67,79^, uniform, or static. Rather, they appear to reflect complex, distributed, and dynamically evolving processes. This aligns with evidence suggesting that CA impacts functional and structural connectivity across large-scale neural networks involved in stress regulation, emotion, and cognition ^76^. Traditional univariate approaches, including voxelwise and ROI-based analyses, may therefore be insufficient to capture the distributed effect. As noted by Popovic et al.^80^, such approaches assume localized specialization and statistical independence, which may not reflect the complexity of the neurobiological signature of CA. Moving forward, adopting data-driven methods that examine multivariate patterns of brain morphology rather than region-specific effects, will be essential to overcome these limitations when investigating associations with specific risk factors.

## Conclusion

Our findings do not support the longstanding hypothesis that CA is associated with reduced hippocampal or amygdala volume in individuals with PD and BD. In contrast, more consistent evidence emerged for reduced GMV in prefrontal regions, suggesting that the neurobiological impact of CA may be more robustly captured within cortical regions.

These results challenge classical limbic-focused models of stress-related neurotoxicity and suggest that the impact of CA may be more diffusely represented across distributed brain networks rather than localized structures. Furthermore, they raise the possibility that conventional structural imaging measures may be too coarse to capture the complex, multilevel processes linking early-life stress to brain outcomes. Future research should prioritize longitudinal designs, refined characterization of trauma (including timing and subtype), systematic consideration of sex differences, and the use of multivariate, data-driven and network-based neuroimaging approaches to better elucidate the neurobiological consequences of CA.

## Data Availability

All data produced in the present work are contained in the manuscript

## ACKNOWLEDGEMENTS

All authors have contributed to the manuscript.

