## Supplementary material for "Revisiting the link between childhood adversity and stress-sensitive brain regions in psychosis and bipolar disorder: A systematic review and meta-analysis"

**Supplementary Table 1.** PRISMA Checklist

| Section/Topic | # | Checklist Item | Page |
|---|---|---|---|
| **TITLE** | | | |
| Title | 1 | Identify the report as a systematic review, meta-analysis or both. | 1 |
| **ABSTRACT** | | | |
| Structured Summary | 2 | Provide a structured summary including as applicable: background; objectives; data sources; study eligibility criteria, participants, and interventions; study appraisal and synthesis methods; results; limitations; conclusions and implications of key findings; systematic review and registration number. | 2 |
| **INTRODUCTION** | | | |
| Rationale | 3 | Describe the rationale for the review in the context of existing knowledge. | 3-4 |
| Objectives | 4 | Provide an explicit statement of objective(s) or question(s) the review addresses with reference to participants, interventions, comparison, outcomes, and study design (PICOS). | 5 |
| **METHODS** | | | |
| Eligibility Criteria | 5 | Specify the inclusion and exclusion criteria for the review and how studies were grouped for syntheses. | 6-7 |
| Information Sources | 6 | Specify all databases, registers, websites, organisations, reference lists and other sources searched or consulted to identify studies. Specify the date when each source was last searched or consulted. | 5 |

**Supplementary material**

| | | | |
|---|---|---|---|
| Search Strategy | 7 | Present full electronic search strategy for at least one database, including any limits used, such that it could be repeated. | 5 and Supplementary material (SM) |
| Selection Process | 8 | State the process for selecting studies (i.e. screening, eligibility, included in systematic review, and if applicable, included in meta-analysis). | 6-7 |
| Data Collection Process | 9 | Specify the methods used to collect data from reports, including how many reviewers collected data from each report, whether they worked independently, any processes for obtaining or confirming data from study investigators, and if applicable, details of automation tools used in the process. | 6 |
| Data Items | 10a | List and define all outcomes for which data were sought. Specify whether all results that were compatible with each outcome domain in each study were sought (e.g. for all measures, time points, analyses), and if not, the methods used to decide which results to collect. | 6 and SM |
| | 10b | List and define all other variables for which data were sought (e.g. participant and intervention characteristics, funding sources). Describe any assumptions made about any missing or unclear information. | 6 and SM |
| Study Risk of Bias Assessment | 11 | Specify the methods used to assess risk of bias in the included studies, including details of the tools used, how many reviewers assessed each study and whether they worked independently, and if applicable, details of automation tools used in the process. | 7 |
| Effect Measures | 12 | Specify for each outcome the effect measures (e.g. risk ratio, mean difference) used in the synthesis or presentation of results. | - |

**Supplementary material**

| | | | |
|---|---|---|---|
| Synthesis Methods | 13a | Describe the processes used to decide which studies were eligible for each synthesis, (e.g. tabulating the study intervention characteristics and comparing against the planned groups for synthesis). | 5-7 |
| | 13b | Describe any methods required to prepare the data for presentation or synthesis, such as handling of missing summary statistics, or data conversion. | 5-7 |
| | 13c | Describe any methods used to tabulate or visually display results of individual studies and synthesis. | 5-7 |
| | 13d | Describe any methods used to synthesise results and provide rationale for the choices. If meta-analysis was performed, describe the models, methods to identify the presence and extent of statistical heterogeneity, and software packages used. | 5-7 |
| | 13e | Describe any methods used to explore possible causes of heterogeneity among study results (e.g. subgroup analyses, meta-regression). | - |
| | 13f | Describe any sensitivity analyses conducted to assess robustness of the synthesised results. | - |
| Reporting Bias Assessment | 14 | Describe any methods used to assess risk of bias due to missing results in a synthesis (arising from reporting biases). | - |
| Certainty Assessment | 15 | Describe any methods used to assess certainty (or confidence) in the body of evidence for an outcome. | 5-7 |
| **RESULTS** | | | |
| Study Selection | 16a | Describe the results of the search and selection process, from the number of records identified in the search to the number of studies included in the review, ideally using a flow diagram. | 7-8 |

**Supplementary material**

| | | | |
|---|---|---|---|
| | 16b | Cite studies that might appear to meet inclusion criteria, but which were excluded, and explain why they were excluded. | 7-9 |
| Study Characteristics | 17 | Cite each included study and present its characteristics. | 7-9 |
| Risk of Bias In Studies | 18 | Present assessment of risk of bias for each included study. | 8 |
| Results in Individual Studies | 19 | For all outcomes, present, for each study: (a) summary statistics for each group (where appropriate) and (b) an effect estimates and its precision (e.g. confidence/credible intervals), ideally using structured tables and plots. | 7-9 |
| Results of Synthesis | 20a | For each synthesis, briefly summarise the characteristics and risk of bias among contributed studies. | 8 |
| | 20b | Present results of all statistical syntheses conducted. If meta-analyses was done, present for each the summary estimate and its precision (e.g. confidence/credible interval) and measures of statistical heterogeneity. If comparing groups, describe the direction of the effect. | - |
| | 20c | Present results of all investigations of possible causes of heterogeneity among study results. | - |
| | 20d | Present results of all sensitivity analyses conducted to assess the robustness of the synthesised results. | - |
| Reporting Bias | 21 | Present assessments of risk of bias due to missing results (arising from reporting biases) for each synthesis assessed. | 7-9 |
| Certainty of Evidence | 22 | Present assessments of certainty (or confidence) in the body of evidence for each outcome assessed. | - |
| **DISCUSSION** | | | |

**Supplementary material**

| | | | |
|---|---|---|---|
| Discussion | 23a | Provide a general interpretation of the results in the context of other evidence. | 10 |
| | 23b | Discuss any limitations of the evidence included in the review. | 15-17 |
| | 23c | Discuss any limitation of the review processes used. | 15-17 |
| | 23d | Discuss implications of the results for practice, policy, and future research. | 17-18 |
| **OTHER INFORMATION** | | | |
| Registration and protocol | 24a | Describe sources of funding for the systematic review and other support; role of funders for the systematic review. | MRC |
| | 24b | Indicate where the review protocol can be accessed, or state that a protocol was not prepared. | Prospero. CRD42022351133 |
| | 24c | Describe and explain any amendments to information provided at registration or in the protocol. | NA |
| Support | 25 | Describe sources of financial or non-financial support for the review, and the role of the funders or sponsors in the review. | NA |
| Competing Interests | 26 | Declare any competing interests of review authors. | NA |
| Availability of Data, Code, and Other Materials | 27 | Report which of the following are publicly available and where they can be found: template data collection forms; data extracted from included studies; data used for all analyses; analytic code; any other materials used in the review. | NA |

*Note.* Adapted from the PRISMA 2020 Statement (Page et al., 2021).

**Supplementary material**

**Supplementary Table 2.** Full List of Search Terms Used in the Literature Search

---

The search term used on OVID:
Mesh terms (starting with “exp” and key words (ending with “mp”) related to CA
exp Sex Offenses/
exp Physical Abuse/
exp Child Abuse/
exp Bullying/
exp Sexual Abuse/
exp Physical Abuse/
exp Emotional Abuse/
exp Child Abuse/
exp CHILD NEGLECT/
exp Emotional Trauma/
exp BULLYING/
exp Abandonment/
exp RAPE/
exp Domestic Violence/
exp Sexual Harassment/
exp Rape/
exp Domestic Violence/
exp Sexual Harassment/
exp sexual abuse/
exp physical abuse/
exp emotional abuse/
exp neglect/
exp child abuse/
exp bullying/
exp sexual bullying/

**Supplementary material**

exp rape/
exp domestic violence/
exp psychotrauma/
exp early life stress/
exp sexual harassment/
(separat* adj5 parent*).mp. [mp=ti, ab, hw, tn, ot, dm, mf, dv, kf, fx, dq, nm, ox, px, rx, ui, sy, tc, id, tm]
victimi*.mp.
(advers* adj5 experienc*).mp. [mp=ti, ab, hw, tn, ot, dm, mf, dv, kf, fx, dq, nm, ox, px, rx, ui, sy, tc, id, tm]
adversit*.mp.
emotional abuse.mp. or exp emotional abuse/
psychological abuse.mp.
neglect.mp.
bully*.mp.
bullied*.mp.
parental loss.mp. or exp parental deprivation/
(Childhood adj5 trauma).mp. [mp=ti, ab, hw, tn, ot, dm, mf, dv, kf, fx, dq, nm, ox, px, rx, ui, sy, tc, id, tm]
(abandon adj10 parent*).mp. [mp=ti, ab, hw, tn, ot, dm, mf, dv, kf, fx, dq, nm, ox, px, rx, ui, sy, tc, id, tm]
maltreat*.mp.
(Parent adj5 loss).mp. [mp=ti, ab, hw, tn, ot, dm, mf, dv, kf, fx, dq, nm, ox, px, rx, ui, sy, tc, id, tm]
(Childhood adj5 maltreat*).mp. [mp=ti, ab, hw, tn, ot, dm, mf, dv, kf, fx, dq, nm, ox, px, rx, ui, sy, tc, id, tm]
early adversity.mp.
being taken into care.mp.
early life stress.mp. or exp early life stress/
exp rape/ or rape.mp.
domestic violence.mp.
sexual harassment.mp.
exp psychotrauma/
sexual abuse.mp.
physical abuse.mp.
Mesh terms (starting with “exp” and key words (ending with “mp”) related to Psychotic disorders and bipolar disorders.

**Supplementary material**

exp psychosis/
exp schizophrenia/
exp schizoaffective psychosis/
exp Psychotic Disorders/
exp Schizophrenia/
exp PSYCHOSIS/
exp Acute Psychosis/
exp Affective Psychosis/
exp SCHIZOPHRENIA/
exp SCHIZOAFFECTIVE DISORDER/
psychot*.mp.
schizophr*.mp.
schizoaf*.mp. [mp=ti, ab, hw, tn, ot, dm, mf, dv, kf, fx, dq, nm, ox, px, rx, ui, sy, tc, id, tm]
hallucinat*.mp.
delusion*.mp.
exp bipolar disorder/
exp manic psychosis/
exp mania/
exp bipolar mania/
exp Bipolar Disorder/
exp Bipolar I Disorder/
exp Bipolar II Disorder/
Bipolar disorder.mp.
mania.mp.
maniaco depressive.mp.
exp Hypomania/
hypomania.mp.
Mesh terms (starting with “exp” and key words (ending with “mp”) related to brain imaging data
exp Neuroimaging/
exp Magnetic Resonance Imaging/

**Supplementary material**

exp White Matter/
exp neuroimaging/
exp imaging/
exp diffusion tensor imaging/
exp nuclear magnetic resonance/
brain scan.mp.
imaging data.mp.
morphometry.mp.
DTI.mp.
neuroimaging.mp.
structural imaging.mp.
exp Gray Matter/
gray matter.mp.
brain volume.mp.
brain size.mp.
brain structure.mp.
exp Magnetic Resonance Imaging/
MRI.mp.
grey matter.mp.

**Supplementary material**

**Supplementary table 3.** Diagnostic codes

| Diagnosis | Code used within ICD-10 | Code used within DSM-IV |
|---|---|---|
| **Schizophrenia** | F20 | 295.10/295.20/295.30/295.60/295.90 |
| **Brief psychotic disorder** | F23 | - |
| **Schizophreniform disorder** | F20.81 | Schizophreniform disorder |
| **Schizoaffective disorder** | F25.0 | 295.70 |
| **Major depressive disorder with psychotic features** | F33.0 | 296.24/296.34 |
| **Psychosis not otherwise specified** | F29 | 298.9 |
| **Bipolar disorder type II** | F31.81 | 296.89 |
| **Bipolar disorder type I (bipolar affective disorder)** | F31.1 | 296.xx (all subtypes) including F31.2 severe with psychotic features |

**Supplementary material**

**Supplementary Table 4.** Newcastle Ottawa Scale

| |
|---|
| **Newcastle Ottawa Scale (adapted from the cohort studies NOS scale)**<br>Note: A study can be awarded a maximum of one star for each numbered item within the Selection and Outcome categories. A maximum of two stars can be given for Comparability. |
| **Selection**<br>1) Representativeness of the exposed cohort (b)<br>a) truly representative of the average individuals with psychosis or bipolar in the community (Really when ALL cases were part of the study - birth cohorts, totally representative) *<br>b) somewhat representative of the average individuals with psychosis or attenuated psychotic symptoms in the community *<br>c) selected group of users eg nurses, volunteers (not applicable for us because of selection criteria excludes these two options)<br>d) no description of the derivation of the cohort (not applicable for us because of selection criteria excludes these two options)<br>2) Selection of the non-exposed cohort (a)<br>a) drawn from the same community as the exposed cohort * (always a, but check that the trauma and non-trauma are extracted from the same cohort – we are not picking traumatised from Norway and non traumatised from Lausanne)<br>b) drawn from a different source (if they were taking this from a different population)<br>c) no description of the derivation of the non-exposed cohort (not applicable for us because of selection criteria excludes these two options)<br>3) Ascertainment of exposure (c) (between b and c for us)<br>a) secure record*<br>b) structured interview*<br>c) written self-report* ( this has been modified: star included here given the common use of self-reports in the field of adversity in psychosis)<br>d) no description (these are excluded)<br>4) Demonstration that outcome of interest was not present at start of study (is the problem already there (the brain abnormality) when the patients have been assessed for the first time? For example, covid infection in pregnant women and we want to see if later children develop autism, the study starts at birth (when the outcome of interest is not present, so it would be "yes", however for most of our clinical samples the problem (low functioning), is already there, so it will likely be "no" but exclude that samples come from prospective studies from birth).<br>a) yes *<br>b) no |

**Supplementary material**

**Comparability (count 2 stars)**

1) Comparability of cohorts on the basis of the design or analysis

a) study controls for confounders * (one star: studies providing adjusted and unadjusted results and/or adjusting by covariates such as age, gender, race, education – often in linear regressions, logistic regressions etc. Correlation, t- test, qui square does not adjust – check statistics; b) study includes a sample of N>=100) *

---

**Outcome**

1) Assessment of outcome

a) independent blind assessment and using validated advances programs * (in imaging data usually this is done by technicians that don't know the traumatic history, programs like Freesurfer)

b) record linkage or formal interview of symptoms (doe sot apply)*

c) self-report

d) no description

2) Was follow-up long enough for outcomes to occur

a) yes * (this will only apply for birth cohorts, for cross-sectional studies does not apply so no star)

b) no

3) Adequacy of follow up of cohorts (it could be that some that drop out were treatment resistant) (>20%)

a) complete follow up (no drop outs) - all subjects accounted for *

b) subjects lost to follow up unlikely to introduce bias - small number lost < 20 % *

c) follow up rate < 80% and no description of those lost

d) no statement/ no follow up (cross sectional studies)

---

**Supplementary material**

**Supplementary Table 4a.** Newcastle Ottawa Scale (NOS) Quality Assessment for the included studies

| | **Psychosis sample** | | | | | | | | | | | |
|---|---|---|---|---|---|---|---|---|---|---|---|---|
| **Author** | **Selection** | | | | | **Comparability** | | **Outcome** | | | | **Total/ Quality Classification** |
| | 1. Represen-tativeness of exposed cohort | 2. Selection of the non-exposed cohort | 3. Ascertain-ment of exposure | 4. Outcome of interest was not present at start of study | Domain Score (out of 4) | 1. Compara-bility of cohorts on the basis of the design or analysis | Domain Score (out of 2) | 1. Assess-ment of outcome | 2. Ade-quate length of follow-up | 3. Adequate follow-up of cohorts | Domain Score (out of 3) | |
| Aas et al., (2012) | * | * | * | | 3/4 | * | 1/2 | * | | | 1/3 | 5/ Fair |
| Alameda et al., (2018) | * | * | * | | 3/4 | ** | 2/2 | * | | | 1/3 | 6/ Good |
| Armio et al., (2020) | * | * | * | | 3/4 | * | 1/2 | * | | | 1/3 | 5/ Fair |
| Begemann et al., (2021) | * | * | * | | 3/4 | ** | 2/2 | * | | | 1/3 | 6/ Good |
| Benedetti et al., (2011) | * | * | * | | 3/4 | * | 1/2 | * | | | 1/3 | 5/ Fair |
| Cancel et al., (2015) | * | * | * | | 3/4 | * | 1/2 | * | | | 1/3 | 5/ Fair |
| Cao et al. (2023) | * | * | * | | 3/4 | ** | 1/2 | * | | | 1/3 | 6/ Good |
| Colic et al. (2024) | * | * | * | | 3/4 | ** | 2/2 | * | | | 1/3 | 6/ Good |
| Del Re et al., (2023) | * | * | * | | 3/4 | ** | 2/2 | * | | | 1/3 | 6/ Good |
| Duarte et al., (2016) | * | * | * | | 3/4 | * | 1/2 | * | | | 1/3 | 5/ Fair |

### Supplementary material

| | | | | | | | | | | | | |
|---|---|---|---|---|---|---|---|---|---|---|---|---|
| Elvsaashagen et al., (2013) | * | * | * | | 3/4 | * | 1/2 | * | | | 1/3 | 5/<br>Fair |
| Du Plessis et al., (2020) | * | * | * | | 3/4 | * | 1/2 | * | | | 1/3 | 5/<br>Fair |
| Fan et al., (2022) | * | * | * | | 3/4 | ** | 2/2 | * | | | 1/3 | 6/<br>Good |
| Frissen et al., (2018) | * | * | * | | 3/4 | * | 1/2 | * | | | 1/3 | 5/<br>Fair |
| Harmata et al., (2023) | * | * | * | | 3/4 | ** | 2/2 | * | | | 1/3 | 6/<br>Good |
| Hernaus et al., (2014) | * | * | * | | 3/4 | * | 1/2 | * | | | 1/3 | 5/<br>Fair |
| Hoffmann et al., (2018) | * | * | * | | 3/4 | ** | 2/2 | * | | | 1/3 | 6/<br>Good |
| Hoy et al., (2012) | * | * | * | | 3/4 | * | 1/2 | * | | | 1/3 | 5/<br>Fair |
| Janiri et al., (2017) | * | * | * | | 3/4 | ** | 22 | * | | | 1/3 | 6/<br>Good |
| Janiri et al., (2017) | * | * | * | | 3/4 | ** | 2/2 | * | | | 1/3 | 6/<br>Good |
| Millman et al., (2022) | * | * | * | | 3/4 | ** | 2/2 | * | | | 1/3 | 6/<br>Good |
| Poletti et al., (2014) | * | * | * | | 3/4 | * | 1/2 | * | | | 1/3 | 5/<br>Fair |
| Quidé et al., (2021) | * | * | * | | 3/4 | ** | 2/2 | * | | | 1/3 | 6/<br>Good |
| Rokita et al., 2020 | * | * | * | | 3/4 | ** | 1/2 | * | | | 1/3 | 5/<br>Fair |
| Ruby et al., (2017) | * | * | * | | 3/4 | | 0/2 | * | | | 1/3 | 4/<br>Poor |
| Sheffield et al., (2013) | * | * | * | | 3/4 | * | 1/2 | * | | | 1/3 | 5/<br>Fair |
| Song et al., (2020) | * | * | * | | 3/4 | * | 1/2 | * | | | 1/3 | 5/<br>Fair |

### Supplementary material

| | | | | | | | | | | | | |
|---|---|---|---|---|---|---|---|---|---|---|---|---|
| Souza-Queiroz et al., (2016) | * | * | * | | 3/4 | * | 1/2 | * | | | 1/3 | 5/ Fair |
| Tseng et al., (2021) | * | * | * | | 3/4 | * | 1/2 | * | | | 1/3 | 5/ Fair |

### Supplementary material

**Supplementary Table 5.** Study Characteristics and Neuroimaging Outcome Parameters

| Author (Year), Country | Diagnosis specification | Diagnosis Measure | CA Variable type | Regions reported in relation to CA | Details on the association of CA on Brain Measurements | Covariates | Inclusion meta-analysis |
|---|---|---|---|---|---|---|---|
| **Aas et al. (2012)** | FEP | ICD-10 | CECA.Q<br>Composite<br><br>(categorical) | 1.Hippocampus<br>2. Amygdala | **1**. NSA (total, left, right)<br>**2**. r = - .40, *p* = - .006<br>Right amygdala: f = 4.80; p = .01<br>Left amygdala: f=4.90; p=.01 | Age<br>Sex<br>ICV | yes |
| **Alameda et al. (2018)** | FEP | DSM-IV | Psychiatric Interview<br>Composite<br>(binary) | 1. Hippocampus<br>2. Amygdala | **1**. NSA Mean and right hippocampus<br>Left hippocampus: d = 0.51, p = 0.046<br>**2.** NSA mean, right and left amygdala | - | no |
| **Armio et al. (2020)** | FEP | DSM-IV | TAD<br>Composite<br><br>(continuous) | Amygdala subnuclei:<br>1. Lateral nucleus<br>2. Basal nucleus<br>3. Cental nucleus<br>4. Accessory basal nucleus<br>5. Corticoamygdaloid transition area | **1.** β = -1.45, t (62) = - 2.852, *p* = .006<br>**2.** β = -0.86, 7 (62) = -2.350, p = 0.022<br>3. NSA<br>4.NSA<br>5. NSA | Age<br>Sex<br>ICV<br>FDR | no |
| **Begemann et al. (2021)** | SCZ<br>BD-I | DSM-IV | CTQ<br>Composite &<br>Subtypes: SA, PA, EA, PN, EN<br>(binary and continuous) | 1.Hippocampus<br>2.Amygdala<br>**68 regions grouped into cortical lobes:**<br>3.Frontal Lobe<br>4.Temporal lobe<br>5.Cingulate/insular lobe<br>6.Subcortical lobe<br>7.Parietal lobe<br>8.Occipital lobe<br>9.Total GMV | **1.** NSA (left and right)<br>**2**. NSA (left and right)<br>**3**. Right medialorbitofrontal region (β = -0.107, p = 0.002)<br>Right paracentral region (β = -0.088, p = 0.018)<br>Right superior frontal area (β = -0.058, p = 0.027)<br>Left precentral regions (β = -0.057, p = 0.0498)<br>**4-9.** NSA<br>NB. Reported results are only significant in the total sample, not significants in PD only. | Age<br>Sex<br>Group<br>Brain volume<br>FDR<br>Type of Scan<br>Medication | no |

### Supplementary material

| | | | | | | | |
|---|---|---|---|---|---|---|---|
| **Benedetti et al. (2011)** | SCZ | DSM-IV (SCID-I) SCZ = 100 % | RFQ<br><br>(binary) | 1.PFC<br>2.ACC<br>3. Hippocampus<br>4. Amygdala | 1. p<0.001<br>2. p<0.001<br>3. NSA<br>4. NSA | Medication<br>FDR correction | no |
| **Cancel et al. (2015)** | SCZ | DSM-IV SCZ = 100% | CTQ Composite & Subtypes : EN, PN, EA, SA<br><br>(continuous) | 1. Total GMV<br>2. Right DLPFC GMV<br>3. Left DLPFC GMV<br>4. Right Inferior Frontal Gyrus | **1.**EN : β = -.47, *p* = .005 ; PA, PN, SA : NSA<br>**2.**EN: Right (k = 1113 voxels, $P_{cluster}$ = 0.003, FWE-corrected, $P_{voxel}$<0.002 FWE-uncorrected, z-score$_{voxels}$ = 4.33 at [40;38;25])<br>**3.**EN: NSA<br>**4.** EN: NSA | Age<br>Gender<br>DUI<br>Parents educational<br>Medication use<br>Bonferroni correction | no |
| **Cao et al. (2024)** | BD-II | DSM-V 100 = BD-II | CTQ (total, PN, EN, SA, EA | 1.Right MFG<br>2.Right orbital MFG | 1.SA, r = -0.348, q<0.01<br>2.PN, r=0.254, q = .03 | Age<br>Sex<br>Education level<br>Medication<br>FDR correction | no |
| **Colic et al. (2022)** | BD | CTQ composite & subtypes : SA, PA, EA, PN, EN (continuous) | sMRI (3T)<br>ROI and whole-brain Automated segmentation | 1.Left hippocampus<br>2.Right hippocampus<br>3.Prefrontal cortex regions surface area (superior frontal, rostral middle frontal, frontal pole, rostral anterior congulate, medial orbitofrontal) | 1.B = -7.41, p = .03 (SA, in women)<br>2.NSA<br>3.Frontal pole surface (CTQ composite, EN, EA, PN in men)<br>Right superior frontal surface area (EN, PA, SA in men) | Age<br>ICV<br>FDR correction | no |
| **Del Re et al. (2023)** | SCZ | DSM-IV-TR | CTQ Composite<br><br>(continuous) | 1.Hippocampus | **1.**Hippocampus<br>Right anterior hippocampus (r = -0.11, p = 0.011)<br>Right posterior hippocampus (r = -0.12, p = 0.003)<br>Left anterior hippocampus NSA<br>Left posterior hippocampus (r =-0.14, p = 0.0004) | Age<br>Sex<br>ICV<br>Race<br>MRI site<br>FDR | no |

### Supplementary material

| | | | | | | | |
|---|---|---|---|---|---|---|---|
| **Duarte et al. (2016)** | BD-I | MINI-Plus | CTQ composite, Subtypes (PN, EN, PA) (continuous) | GMV:<br>1.Right DLPFC<br>2.Bilateral Thalamus | **1.** CTQ Total : Z=4.37, voxel= 149, p=.013 ; PA : Z=4.04, voxel= 28, p=.04<br>**2.** CTQ Total : Z=3.54, voxel= 41, p=.021<br>PN : Right Thalamus, Z=4.25, voxel= 321, p=.002<br>Left thalamus Z=3.30, voxel=15, p=.04<br>EN: Right thalamus Z=3.31, voxel=29, p=.04<br>(All results show a negative association) | Age<br>Sex<br>Total GMV | no |
| **Du Plessis et al. (2020)** | FEP | DSM-IV | CTQ<br>Composite<br>(continuous) | 1.Hippocampus | **1.** NSA | Age<br>Sex<br>Diagnosis<br>Scanner-Sequence<br>ICV<br>Substance Abuse | yes |
| **Elvsåshagen et al. (2013)** | BD II | DSM-IV | CTQ composite<br>(continuous) | Hippocampus subfields<br>1.Left DG–CA4<br>2. Right DG–CA4<br>3. Left CA2–3 | 1. NSA<br>2. NSA<br>3. NSA | Total GMV<br>Age<br>Group status | no |
| **Fan et al. (2022)** | SCZ | DSM-IV | CTQ composite and subtypes<br><br>(continuous) | 1.Amygdala<br>2.Thalamus<br>3.Caudate<br>4.Putamen<br>5.Pallidum<br>5.Nucleus accumbens<br>6.Hippocampus<br>7.Total cortical GMV | 1.r =-0.20, p = 0.03<br>2.NSA<br>3.NSA<br>4.NSA<br>5.NSA<br>6.NSA<br>7.NSA | Age<br>Sex<br>ICV<br>Bonferroni correction | yes |
| **Frissen et al. (2018)** | SCZ | DSM-IV | CTQ<br>Composite<br><br>(continuous) | 1.Total GMV | **1.**B = -9.79, *p* = .01 | Age<br>Sex<br>Educational level<br>ICV<br>Scan type | no |
| **Harmata et al. (2023)** | BD-I | SCID<br>DSM-V<br>BD-I | ACE score | 1.Left cerebellar GMV<br>2.Right cerebellar GMV<br>3.Vermis I-V | 1.p=0.019, q=0.033<br>2.p=0.018, q=0.033<br>3.NSA | Age<br>Sex<br>ICV | no |

### Supplementary material

| | | | | | | | |
|---|---|---|---|---|---|---|---|
| | | | | 4.Vermis VI-VII<br>5.Vermis VII-X | 4.NSA<br>5.NSA | FDR correction | |
| **Hernaus et al. (2014)** | SCZ | DSM-IV | CTQ<br>Composite<br>(continuous) | 1.Left Hippocampus<br>2.Right Hippocampus | **1.** NSA<br>**2** .NSA | ICV<br>Age<br>Gender<br>Educational level<br>Drug use | yes |
| **Hoffmann et al. (2018)** | SCZ<br>SZA | DSM-IV | CAQ<br>Composite &<br>Subtypes: PA, SA, EN, EA, loss, family dysfunction, financial dysfunction<br>(continuous) | 1. Caudate<br>2. Putamen<br>3. Nucleus Accumbens<br>4. DLPFC<br>5.Hippocampus | **1.** Right Caudate, PA: B =16.00, 95% CI = 44.00-26.00, p = 0.008.<br>**2**. NSA<br>**3.** NSA<br>**4.** NSA<br>**5**. NSA | Age<br>Gender<br>Scanning site<br>ICV | no |
| **Hoy et al. (2011)** | FEP | ICD 10 | TEC<br>Composite<br>(Binary) | 1. Total GMV, WM, CFS & ICV<br>2. Hippocampus<br>3. Amygdala<br>4. Hippocampus & Amygdala Complex | **1.** NSA<br>**2.** NSA<br>Right hippocampus: NSA<br>Left hippocampus: β = -.40, *p* =.03<br>**3.** β = -0.5, p = 0.019<br>Right Amygdala: β = -.42, p = .04<br>Left Amygdala: NSA<br>**4.** β = .45, *p* = .03 | Age<br>Sex<br>Delay to scan time | yes |
| **Janiri et al., (2017)** | BD-I<br>BD-II | DSM-IV<br>BD-I<br>BD-II | CTQ<br>Composite<br>(continuous) | 1. Amygdala<br>2.Hippocampus | 1.Right (BD-I) (t=2.88, p=.006)<br>Right (BD-II) NSA<br>Left (BD-I) (t=1.54, p=.012)<br>Left (BD-II) (2.54, p=.01)<br>2.Right (BD-I) NSA<br>Right (BD-II) (t=2.27, p=.02)<br>Left (BD-I) NSA<br>Left (BD-II) (t=2.54, p=.01)<br>(results indicate bigger volume in patients in relation to CA) | ICV<br>Sex | yes |
| **Janiri et al., (2019)** | BD-I<br>BD-II | DSM-IV | CTQ<br>Composite<br>(continuous) | Hippocampal subfield volume<br>1.Right CA1 | **1.**t=1.12, .26 (BD-I)<br>t=1.70, p=.09 (BD-II)<br>**2.** t=2.28, .02 (BD-I) | DUI | no |

### Supplementary material

| | | | | | | | |
|---|---|---|---|---|---|---|---|
| | | | | 2. Right Presubiculum<br>3. Right subiculum<br>4.Left CA1<br>5. Left Presubiculum<br>6.Left subiculum | t=3.11, p=.003 (BD-II)<br>**3.** t=2.28, .02 (BD-I)<br>t=3.11, p=.003 (BD-II)<br>**4.**t=.66, .50 (BD-I)<br>t=2.35, p=.002 (BD-II)<br>**5.**t=1.65, .10 (BD-I)<br>t=3.37, p=.001 (BD-II)<br>**6.**t=2.02 .04 (BD-I)<br>t=2.75, p=.008 (BD-II)<br>(results indicate bigger volume in relation to CA) | | |
| **Millman et al. (2022)** | SCZ | DSM-IV-TR (SCID) | CTQ | 1.Left and right amygdala<br>2.Left and right hippocampus | **1.**Left amygdala: r = 0.30, p=0.01<br>Right amydala NSA<br>**2.**Left hippocampus: NSA<br>Right hippocampus: NSA | Sex<br>ICV | yes |
| **Poletti et al., (2014)** | BD-I | DSM-IV | RFQ<br>Composite<br>(continuous) | | **1.** MNI coordinates −33 −15 -21 ; T = 3.53, Z = 3.38, p FWE = .02<br>(results indicate a volume reduction in relation to CA) | Yes<br>Age<br>Sex<br>DUI<br>No.of eps.<br>Handedness | no |
| **Quidé et al. (2021)** | SCZ<br>SZA<br>BD | ICD-10 | CTQ<br>Composite (continuous),<br>High, average and low trauma exposure | 1. IC2:<br>Precuneus, Cingulate Gyrus, Inferior Parietal Lobule, Posterior Cingulate, Superior Parietal Lobule<br>2.IC6:<br>Declive, Tuber, Lentiform Nucleus, Culmen, Uvula, Cerebellar Tonsil, Pyramis, Caudate, Fusiform Gyrus | SCZ, SZA<br>**1**. t = 2.064, p = .041 (low trauma)<br>**2**. NSA<br><br>BD<br>1.NSA<br>2.NSA | Age<br>Sex<br>ICV<br>Group status<br>Bonferroni adjustments | no |
| **Rokita et al. (2020)** | SCZ<br>SZA | DSM-IV | CTQ<br>Composite & PN<br>(Continuous) | 1.Amygdala<br>2.Hippocampus<br>3.ACC | **1**.NSA<br>**2**.NSA<br>**3**.r = -0.161, p = 0.048 | Age<br>Sex<br>Years of Education<br>ICV | no |

### Supplementary material

| | | | | | | | |
|---|---|---|---|---|---|---|---|
| **Ruby et al. (2017)** | SCZ | DIGS | Early Trauma Inventory Composite (Continuous) | 1.Whole Brain volume<br>2.Amygdala/Whole Brain Volume Ratio<br>3.Hippocampal/Amydala Volume Ratio<br>4.Hippocmapus/Whole Brain Volume Ratio | **1**.r = -.50, (n = 17), *p* = .04<br>**2**.r = .56, (n = 17), *p* = .02<br>**3**.NSA<br>**4**.NSA | - | yes |
| **Sheffield et al. (2013)** | SCZ<br>SZA<br>BP-I | DSM-IV | CTQ Composite & Subtypes: PA, SA, EA, EN, PN (Binary) | 1.total GMV<br>2. 7 regions within the PFC<br>3. Hippocampus<br>4. Amygdala | **1**.Total CTQ (r = -.27, *p* = .04)<br>SA (r = -.34, *p* = .008)<br>NSA PA,EA, EN, PN,<br>**2**. Left middle frontal gyrus (k = 265, -48, 23, 31), p < 0.005<br>**3.** NSA<br>**4.** NSA | Age<br>Sex | no |
| **Song et al., (2020)** | BD-I<br>BD-II | DSM-V | CTQ Composite (Continuous) | 1.Right precentral gyrus<br>2.Left Middle Frontal Gyrus | 1.r=-.61, *p*<.0003<br>2.NSA | Age<br>Sex<br>ICV | no |
| **Souza-Queiroz et al., (2016)** | BD | DSM-IV | CTQ Composite Subtypes (EN/PN) (continuous) | 1.Hippocampus<br>2.Amygdala | **1.** NSA<br>**2**. β = -0.22, p = .04 (HC and BD), BD only NSA | Yes<br>Age<br>Gender<br>Education<br>ICV<br>DUI<br>Medication | no |
| **Tseng et al. (2021)** | SCZ | DSM-IV | BBTS Composite (Continuous) | 1. Thalamus<br>2. Inferior Fontal Gyrus<br>3. Insula<br>4. Superior Frontal Gyrus<br>5. Precuneus | **1.**NSA<br>**2.** NSA<br>**3.** NSA<br>**4.** NSA<br>**5.** NSA | DUI<br>Medication | no |

**Supplementary material**

**Supplementary table 6.** Summary of meta-regressions

| Meta-analysis | Hemisphere | Moderator | β coefficient | 95% CI | p-value | Heterogeneity ($I^2$) | Between study variance ($\tau^2$) |
|---|---|---|---|---|---|---|---|
| Hippocampus | Left | Age | 0.010 | -0.016; 0.037 | 0.449 | 51.48% | 0.021 |
| | | Sex | 0.002 | -0.020; 0.023 | 0.871 | 62.26% | 0.026 |
| | | Illness stage | 0.061 | -0.297; 0.419 | 0.738 | 55.45% | 0.022 |
| | | Segmentation Method | -0.081 | -0.487; 0.325 | 0.696 | 62.7% | 0.025 |
| | | Study Quality (NOS) | 0.217 | -0.055; 0.491 | 0.118 | 29.43% | 0.007 |
| | Right | Age | 0.008 | -0.016; 0.033 | 0.495 | 46.11% | 0.016 |
| | | Sex | -0.004 | -0.023; 0.0157 | 0.708 | 49.95% | 0.015 |
| | | Illness stage | 0.202 | -0.042; 0.445 | 0.105 | 41.46% | 0.012 |
| | | Segmentation Method | -0.049 | -0.428; 0.330 | 0.801 | 41.46% | 0.019 |
| | | Study Quality (NOS) | 0.242 | 0.023; 0.461 | **0.030** | 41.46% | 0.011 |
| | Total | Age | 0.011 | -0.016; 0.038 | 0.443 | 60.16% | 0.031 |
| | | Sex | 0.020 | 0.007; 0.035 | **0.007** | 7.31% | 0.001 |
| | | Illness stage | 0.125 | -0.271; 0.520 | 0.537 | 59.77% | 0.023 |
| | | Segmentation Method | -0.281 | -0.583; 0.022 | 0.068 | 42.63% | 0.014 |
| | | Study Quality (NOS) | 0.230 | 0.042; 0.418 | **0.016** | 25.60% | 0.006 |
| Amygdala | Left | Age | 0.016 | -0.046; 0.078 | 0.614 | 85.5% | 0.069 |
| | | Sex | -0.013 | 0.026; 0.037 | 0.611 | 85.83% | 0.102 |
| | | Illness stage | 0.513 | 0.199; 0.122 | **0.01** | 47.66% | 0.016 |
| | | Segmentation method | -0.516 | -0.903; -0.991 | **0.01** | 47.66% | 0.016 |
| | | Study Quality (NOS) | 0.513 | 0.199; 0.122 | **0.01** | 47.66% | 0.016 |
| | Right | Age | 0.018 | -0.017; 0.054 | 0.318 | 60.09% | 0.036 |
| | | Sex | -0.005 | -0.044; 0.034 | 0.798 | 73.12% | 0.046 |
| | | Illness stage | 0.378 | 0.085; 0.671 | **0.011** | 0% | 0.000 |
| | | Segmentation method | -0.378 | -0.671; -0.085 | **0.011** | 0% | 0.000 |
| | | Study Quality (NOS) | 0.378 | 0.08; 0.671 | **0.011** | 0% | 0.000 |
| | Total | Age | 0.010 | -0.039; 0.059 | 0.689 | 85.76% | 0.113 |
| | | Sex | 0.001 | -0.044; 0.046 | 0.958 | 83.41% | 0.090 |
| | | Illness stage | 0.603 | 0.123; 1.083 | **0.014** | 82.58% | 0.082 |
| | | Segmentation method | -0.202 | -0.743; 0.339 | 0.465 | 83.84% | 0.086 |
| | | Study Quality (NOS) | -0.072 | -0.471; 0.327 | 0.098 | 85.92% | 0.098 |

### Supplementary material

#### Supplementary Figure 1a

Forest plot – association between left hippocampal volume and CA

| Study name | Correlation | Lower limit | Upper limit | Z-Value | p-Value |
|---|---|---|---|---|---|
| Aas et al. (2012) | 0.100 | -0.199 | 0.382 | 0.650 | 0.516 |
| Hernaus et al. (2014) | -0.200 | -0.392 | 0.009 | -1.880 | 0.060 |
| Hoy et al. (2011) | -0.400 | -0.709 | 0.038 | -1.797 | 0.072 |
| Janiri et al. (2017) | 0.090 | -0.103 | 0.277 | 0.911 | 0.362 |
| Millman et al. (2022) | 0.060 | -0.165 | 0.279 | 0.520 | 0.603 |
| Pooled | -0.036 | -0.196 | 0.127 | -0.427 | 0.669 |
| Prediction Interval | -0.036 | -0.483 | 0.427 | | |

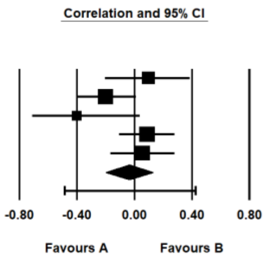


Meta Analysis

#### Supplementary Figure 1b

Forest plot – association between right hippocampal volume and CA

| Study name | Correlation | Lower limit | Upper limit | Z-Value | p-Value |
|---|---|---|---|---|---|
| Aas et al. (2012) | 0.050 | -0.247 | 0.339 | 0.324 | 0.746 |
| Hernaus et al. (2014) | -0.190 | -0.383 | 0.019 | -1.784 | 0.074 |
| Hoy et al. (2011) | -0.160 | -0.553 | 0.292 | -0.685 | 0.494 |
| Janiri et al. (2017) | 0.080 | -0.113 | 0.268 | 0.810 | 0.418 |
| Millman et al. (2022) | 0.180 | -0.044 | 0.387 | 1.576 | 0.115 |
| Pooled | 0.010 | -0.138 | 0.158 | 0.135 | 0.893 |
| Prediction Interval | 0.010 | -0.388 | 0.405 | | |

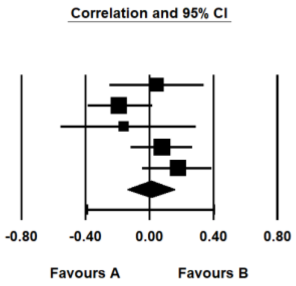


Meta Analysis

**Supplementary material**

### Supplementary Figure 2a

Forest plot – association between left amygdala volume and CA

| Study name | Correlation | Lower limit | Upper limit | Z-Value | p-Value |
|---|---|---|---|---|---|
| Aas et al. (2012) | -0.150 | -0.425 | 0.150 | -0.980 | 0.327 |
| Hoy et al. (2011) | -0.520 | -0.777 | -0.114 | -2.445 | 0.014 |
| Janiri et al. (2017) | 0.120 | -0.073 | 0.305 | 1.218 | 0.223 |
| Millman et al. (2022) | 0.300 | 0.083 | 0.490 | 2.681 | 0.007 |
| Pooled | -0.022 | -0.309 | 0.268 | -0.147 | 0.883 |
| Prediction Interval | -0.022 | -0.869 | 0.858 | | |

Statistics for each study

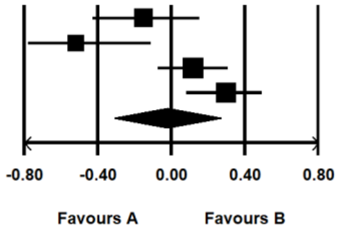


Meta Analysis

### Supplementary Figure 2b

Forest plot – association between right amygdala volume and CA

| Study name | Correlation | Lower limit | Upper limit | Z-Value | p-Value |
|---|---|---|---|---|---|
| Aas et al. (2012) | -0.150 | -0.425 | 0.150 | -0.980 | 0.327 |
| Hoy et al. (2011) | -0.420 | -0.721 | 0.014 | -1.899 | 0.058 |
| Janiri et al. (2017) | 0.150 | -0.043 | 0.332 | 1.526 | 0.127 |
| Millman et al. (2022) | 0.120 | -0.105 | 0.334 | 1.044 | 0.296 |
| Pooled | -0.017 | -0.230 | 0.198 | -0.153 | 0.878 |
| Prediction Interval | -0.017 | -0.709 | 0.692 | | |

Statistics for each study

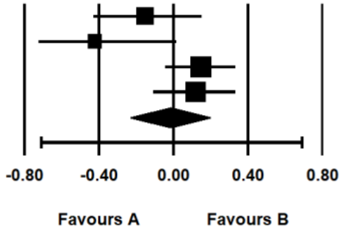


Meta Analysis

### Supplementary material

### Supplementary Figure 3a

Forrest plot – association between total hippocampal volume and CA

| Study name | Correlation | Lower limit | Upper limit | Z-Value | p-Value |
|---|---|---|---|---|---|
| | Statistics for each study | | | | |
| Aas et al. (2012) | -0.150 | -0.425 | 0.150 | -0.980 | 0.327 |
| Fan et al. (2023) | -0.187 | -0.350 | -0.013 | -2.107 | 0.035 |
| Hernaus et al. (2014) | -0.190 | -0.575 | 0.263 | -0.816 | 0.414 |
| Hoy et al. (2011) | -0.315 | -0.657 | 0.135 | -1.383 | 0.167 |
| Janiri et al. (2017) | 0.085 | -0.108 | 0.272 | 0.861 | 0.389 |
| Millman et al. (2022) | 0.120 | -0.105 | 0.334 | 1.044 | 0.296 |
| Ruby et al. (2018) | -0.470 | -0.717 | -0.118 | -2.550 | 0.011 |
| Pooled | -0.118 | -0.271 | 0.040 | -1.462 | 0.144 |
| Prediction Interval | -0.118 | -0.510 | 0.315 | | |

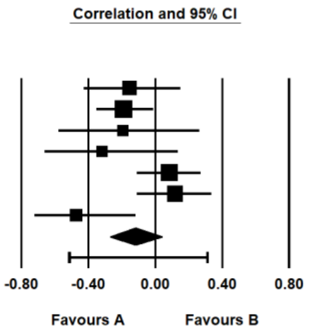


Meta Analysis

### Supplementary Figure 3b

Forrest plot – association between total amygdala volume and CA

| Study name | Correlation | Lower limit | Upper limit | Z-Value | p-Value |
|---|---|---|---|---|---|
| | Statistics for each study | | | | |
| Aas et al. (2012) | -0.400 | -0.621 | -0.121 | -2.746 | 0.006 |
| Fan et al. (2023) | -0.200 | -0.362 | -0.027 | -2.258 | 0.024 |
| Hoy et al. (2011) | -0.500 | -0.766 | -0.087 | -2.331 | 0.020 |
| Janiri et al. (2017) | 0.130 | -0.063 | 0.314 | 1.320 | 0.187 |
| Millman et al. (2022) | 0.210 | -0.013 | 0.413 | 1.846 | 0.065 |
| Ruby et al. (2017) | 0.560 | 0.126 | 0.814 | 2.451 | 0.014 |
| Pooled | -0.042 | -0.296 | 0.218 | -0.310 | 0.756 |
| Prediction Interval | -0.042 | -0.727 | 0.686 | | |

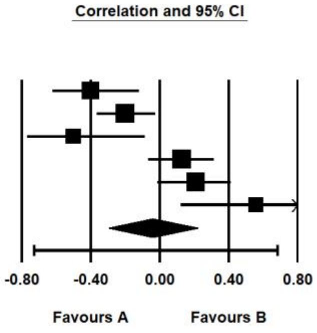


Meta Analysis
